# Cognitive and Mental Health Profiles of Binge-Eating Populations with and without Comorbid Addiction

**DOI:** 10.1101/2024.09.11.24313520

**Authors:** Jake Jeong, Jungwon Jang, Giho Jeon, Kwangyeol Baek

## Abstract

Binge eating disorder (BED) is one of the most prevalent eating disorders and involves an increased risk of mental health problems, obesity and metabolic disease. Recent studies suggest that BED is similar to addictive disorders in its phenomenology and neurobiological mechanisms. Comorbid addiction (e.g., substance use disorders and behavioral addiction) is also very frequent in BED patients. However, it is still unclear whether BED population with comorbid addictions differ in their cognitive and mental health characteristics (e.g., impulsivity, behavioral inhibition, self-control, emotion regulation, mood and anxiety) than those without comorbid addiction. In the present study, we compared various psychometric scales across 30 binge-eating individuals with co-occurring addictive behaviors (i.e., alcohol, nicotine, gambling, and video games), 32 binge-eating individuals without addiction, and 178 healthy control subjects with neither binge-eating nor addiction. Both binge-eating groups showed a significant increase in inhibition motivation (BAS/BIS-inhibition), perceived stress, and state/trait anxiety compared to healthy controls, but there was no difference between the two binge-eating groups. Higher impulsivity and lower self-control were observed in both binge-eating populations to a significantly larger extent in the group with comorbid addiction. Interestingly, significantly increased depression and impaired emotion regulation (less use of cognitive reappraisal) were observed only in the binge-eating group with comorbid addiction when compared to the healthy controls. Our findings demonstrated the commonality and difference in binge-eating populations with and without comorbid addiction. It will help to elucidate cognitive and mental health aspects of comorbid addiction in BED and develop more tailored diagnoses and treatments.

## 1 Introduction

Binge eating is one of the most common behavioral symptoms in eating disorders, particularly binge eating disorder (BED). BED was formally included as a new diagnosis in DSM-5, which is characterized by recurrent episodes of binge eating (at least once a week for 3 months), marked distress regarding binge eating, and absence of regular compensatory behaviors (a major difference from Bulimia Nervosa). A binge-eating episode in BED is characterized by eating a definitely larger amount of food than typical in “a discrete period of time” (e.g., within 2 hours) and “the sense of lack of control over eating during the episode”, which makes it different from just overeating.

BED is the most common eating disorder and usually involves serious mental and physical health issues. BED’s lifetime prevalence is estimated at 1.25% for women and 0.42% for men in the United States (Udo & Grilo, 2018) and 1.9% for women and 0.7% for men in Australia (Bagaric et al., 2020). BED was associated with obesity, type 2 diabetes, hypertension, and elevated cholesterol and triglycerides (Udo & Grilo, 2019). Moreover, BED was significantly associated with mental disorders as well: a lifetime prevalence of another psychiatric disorder was 93.8% in individuals with BED. The most prevalent comorbidity was major depressive disorder (69.9% lifetime prevalence) followed by alcohol use disorder (52.0%), borderline personality disorder (49.2%), nicotine use disorder (40.2%), general anxiety disorder (33.0%), etc. A history of suicide attempts was reported in 22.9% of individuals with BED as well. Despite its detrimental impacts on health and quality of life, people rarely seek treatment for binge eating but ask for medical care for other complications such as morbid obesity, diabetes, metabolic syndromes, and comorbid mental disorders.

The psychopathological and neurobiological underpinnings of binge eating are still under investigation, but recent studies have implicated similarities with addictive disorders. Characteristic behaviors of BED, such as eating larger amounts of food than intended, lack of control, overeating, and binging despite adverse consequences, are comparable to diagnostic symptoms of addictive disorders (or substance use disorders). Animal model studies of binge eating have reported dysfunctional dopaminergic and opioid pathways as well as reduced activity in the prefrontal cortex, which might be a common neurobiological mechanism for binge eating and substance addiction. In animal models, highly palatable foods (high in sugar and/or fat) can increase dopamine levels and strengthen the stimulus-reward association similar to drugs of abuse and induce binge-like consumption behaviors. Human neuroimaging studies also demonstrated alterations in prefrontal and striatal circuits of reward processing and cognitive control in binge-eating and obese subjects.

Despite this evidence of shared neural mechanisms for binge-eating and addictive behaviors, there are few studies of binge-eating individuals in respect of comorbid addictions. Most epidemiological studies of BED reported only the prevalence of comorbid addictive disorders. Peterson et al. (2005) tried subtyping BED into BED subjects with and without substance use disorder (n=39 and 45, respectively), and they found more binge-eating episodes and higher impulsivity traits in BED subjects with substance addiction. In a later study, Becker and Grilo (2015) identified four subgroups in 347 BED patients (34 subjects with co-occurring substance addiction, 129 with mood disorder, 60 with both substance addiction and mood disorders, and 124 with neither of them). In this study, 34 BED subjects with substance addiction were not different from the group with only BED in personality traits, but the groups with comorbid mood disorders showed increased depression symptoms and lower self-esteem. However, these studies are not explicitly designed to contrast binge-eating populations with and without addictive disorders and did not fully assess many cognitive aspects related to addictive behaviors (e.g., motivational system, impulsivity, self-control, emotion regulation, perceived stress, etc.). Another limitation of previous studies is that most of them investigated the comorbidity of ‘substance addiction’ but not ‘behavioral addiction’. Gambling disorder and internet gaming disorder are formally established as behavioral addictions now, so the comorbidity of such addictive behavior should be taken into account.

This study aimed to compare binge-eating individuals with and without co-occurring addictive behaviors to investigate the psychopathological underpinnings in binge-eating populations with comorbid addiction. We also compared both binge-eating populations with the age-matched healthy controls to identify cognitive and mental health characteristics linked to binge-eating. We hypothesized that increased impulsivity and decreased self-control might be associated with binge-eating behaviors, particularly more strongly in individuals with comorbid addictions. We also hypothesized that binge-eating individuals might suffer mental health problems such as increased stress and anxiety, as well as depression and impaired emotion regulation.

## 2 Materials and methods

### 2.1 Participants

This study was conducted with the approval of the Institutional Review Board (IRB) of Pusan National University. We recruited participants from the Korean adult population, ages 19 to 59, who were not diagnosed with any mental disorders except for sleep disorders. Recruitment was conducted using advertisements online, e.g., internet bulletin boards in 7 local universities, mobile apps for the local community, and SNS messengers. A total of 350 participants were initially recruited between May and June 2022. Exclusion criteria were the following: 1) incomplete assessments, 2) inconsistent responses, and 3) positive responses regarding illegal drug use or misuse of psychoactive medications. Thirty-eight subjects were excluded, resulting in a total of 312 participants for data analysis. Among 312 participants, 195 subjects were females (62.5%). For age distribution, there were 196 participants of 29 years old or younger (62.8%), 68 participants in their 30s (21.8%), 29 participants in their 40s (9.3%), and 19 participants in their 50s (6.1%).

### 2.2 Screening Tools for Binge-eating and Addictions

#### 2.2.1 Binge-eating

The Binge Eating Scale (BES) is a widely used 16-item self-administered screening measure of binge eating (Gormally et al., 1982). Each item is assigned a score ranging from 0 to 2 points or 0 to 3 points based on their respective weights. Total scores below 18 points indicate ‘no binge-eating,’ while scores between 18 and 26 points signify ‘moderate level of binge-eating,’ and scores of 27 or higher indicate ‘severe level of binge-eating.’ In this study, scores equal to or exceeding 18 were considered indicative of the presence of BED. The internal consistency (Cronbach’s α) in the Korean validation study was determined to be .84 (Lee and Hyun, 2001).

#### 2.2.2 Addictions

##### Alcohol

The Alcohol Use Disorders Identification Test (AUDIT) was used to assess alcohol addiction (Babor et al., 2001). The scale consists of 10 items, with responses ranging from 0 to 4 points on a 5-point Likert scale. The clinical cut-off for diagnosis is set at 12 points, and scores equal to or above 12 are indicative of alcohol use disorder (AUD). The internal consistency (Cronbach’s α) in the Korean validation study was reported as .92 (Lee, 2000).

##### Nicotine

The Fagerstrom Test for Nicotine Dependence (FTND) was used to examine nicotine dependence (Heatherton et al., 1991). The test consists of 6 items, with two items scored on a scale of 0 to 3 point, and the remaining items using a dichotomous ‘yes/no’ response. In this study, we utilized a clinical cut-off of 7 or higher. The internal consistency (Cronbach’s α) in the Korean validation study was reported as .69 (Ahn et al., 2002).

##### Video Games

The Clinical Video Game Addiction Test 2.0 (C-VAT 2.0) assesses video game addiction (Van Rooij et al., 2017). The test consists of 11 items, using a binary ‘yes (1 point)’ or ‘no (0 points)’ response. A total score of 6 points or higher indicates the presence of video game addiction. The internal consistency (Cronbach’s α) in the Korean validation study was reported as .94(Jang et al., 2020).

##### Gambling

The Canadian Problem Gambling Index (CPGI) measured problem gambling behavior (Ferris and Wynne, 2001). The index comprises 9 items, using a 4-point Likert scale ranging from 0 to 4 points. Scores of 0 indicate ‘non-problem gambling’, scores between 1 and 2 indicate ‘low-risk gambling’, scores between 3 and 7 indicate ‘moderate-risk gambling’, and 8 or higher are categorized as ‘problem gambling’. A cut-off score of 8 or higher was utilized in this study. The internal consistency (Cronbach’s α) in the Korean validation study was reported as .86(Kim et al., 2011).

### 2.3 Clinical questionnaires

#### 2.3.1 Psychometric scales for Cognitive functions

##### Behavioral Inhibition/Activation Systems

The Behavioral Inhibition/Activation System Scale (BIS/BAS) assessed behavioral activation and inhibition (Carver and White, 1994). The scale comprises 20 items, using a 4-point Likert scale ranging from 1 to 4 points. The behavioral inhibition system (BIS) is composed of a single factor, while the behavioral activation system (BAS) consists of three sub-factors representing a strong desire to engage in activities (‘Drive’), seeking novelty and approach behavior toward potential rewards (‘Fun Seeking’), and positive responsiveness to rewards (‘Reward Responsiveness’). Higher scores on each factor indicate a higher propensity for the associated tendencies. The internal consistency (Cronbach’s α) in the Korean validation study was reported as .78 (Kyo Heon and Wuon Shik, 2001).

##### Impulsivity

The Barratt Impulsiveness Scale-11 (BIS-11) was used to quantify impulsivity (Patton et al., 1995). The scale consists of 30 items, using a 4-point Likert scale ranging from 1 to 4 points. Higher total scores on the scale indicate higher levels of impulsivity. The internal consistency (Cronbach’s α) in the Korean validation study was reported as .80 (Heo et al., 2012).

##### Self-Control

The Brief Self-Control Scale (BSCS) measures the degree of self-control (Tangney et al., 2018). The scale comprises 11 items, using a 5-point Likert scale ranging from 1 to 5 points. Higher scores on the scale indicate higher levels of self-control ability. The internal consistency (Cronbach’s α) in the Korean validation study was reported as .81(Hong et al., 2012).

##### Emotion Regulation

The Emotion Regulation Questionnaire (ERQ) was developed to examine emotion regulation (Gross and John, 2003). The scale consists of 10 items, using a 7-point Likert scale ranging from 1 to 7 points. The emotion regulation scale (ERQ) comprises two factors: ‘Reappraisal’, which reflects the cognitive reinterpretation of emotions in response to antecedent events, and ‘Suppression’, which reflects the tendency to inhibit the expression of one’s feelings. Higher scores on each factor indicate a higher propensity for the associated strategy. The reliability (Cronbach’s α) reported in the Korean validation study was .85 for reappraisal and .73 for suppression (Jae-Min, 2005).

#### 2.3.2 Psychometric scales for Mental health

##### Depression

The Patient Health Questionnaire-9 (PHQ-9) was used to quantify depression symptoms (Kroenke et al., 2001). The scale comprises nine items, using a 4-point Likert scale ranging from 0 to 3 points. Total scores ranging from 0 to 4 indicate ‘no depression,’ scores between 5 and 9 indicate ‘mild depression,’ scores between 10 and 19 indicate ‘moderate depression,’ and scores of 20 or higher are classified as ‘severe depression’. The internal consistency (Cronbach’s α) in the Korean validation study was reported as 0.81(Park et al., 2010).

##### Stress

The Perceived Stress Scale (PSS) was used to assess the degree of perceived stress(Cohen et al., 1983). The scale consists of 10 items, using a 5-point Likert scale ranging from 0 to 4 points. Higher scores on the scale indicate a higher level of perceived stress. In the Korean validation study, the reliability (Cronbach’s α) was reported as .74 for positive perception and .77 for negative perception (Park and Seo, 2010).

##### Anxiety

The State/Trait Anxiety Inventory-X (STAI-X) quantified the level of anxiety in each individual (Spielberger, 1970). The inventory comprises 40 items on a 4-point Likert scale, with 20 items each for measuring state and trait anxiety. For state anxiety, total scores of 51 or below indicate ‘normal,’ scores between 52 and 56 indicate ‘mild,’ scores between 57 and 61 indicate ‘moderate,’ and 62 or higher suggest ‘severe.’ For trait anxiety, total scores of 53 or below indicate ‘normal,’ scores between 54 and 58 indicate ‘mild,’ scores between 59 and 63 indicate ‘moderate,’ and 64 or higher suggest ‘severe.’ The reliability (Cronbach’s α) reported in the Korean validation study was .87 for state anxiety and .86 for trait anxiety(Kim and Sin, 1978).

### 2.4 Statistical analysis

Descriptive statistics were calculated to compare the general characteristics of the groups. Initially, group differences in demographic variables (gender, age, BMI) were assessed using the chi-square test and one-way ANOVA. In the remaining data analysis, Analysis of Covariance (ANCOVA) with gender and BMI as covariates was conducted to assess group differences while controlling the effect of gender and BMI. The Bonferroni correction was applied in ANCOVA to address multiple comparisons regarding 12 psychometric scales for cognitive and mental health characteristics. In instances where a significant group difference was identified, post-hoc Tukey’s tests were conducted. All statistical analyses were performed using R 4.3.1 software.

## 3 Results

### 3.1 Demographic information in the Study groups

Depending on the BES and four addiction scales (AUDIT, FTND, C-VAT, CPGI), participants were classified into three groups: 1) the health control (HC) group, 2) the binge-eating only (BE-only) group, and 3) the binge-eating with comorbid addictive disorder (BE+AD) group (as shown in Table 1). The HC group consisted of 178 participants who did not meet any clinical criteria for binge-eating and addiction. The BE-only group comprised 32 participants who met the clinical cut-off for binge eating only. The BE+AD group included 30 participants who satisfied the clinical cut-off values in both binge-eating behavior and at least one of the addiction screening scales. Among 30 subjects in the BE+AD groups, 19 had video game addiction, 16 had alcohol addiction, 9 had gambling disorder, and one subject had nicotine dependence (13 participants had multiple addictions, and 17 had a single type of addiction). The mean number of addictive behaviors in this group was 1.4 (range: 1-3, SD: 0.71). The remaining 72 subjects with only addictive behaviors were not included in the present analysis.

**Table 1.** The demographic information and the addiction profiles of the study groups. Mean (SD) except for the gender. BMI: Body-Mass Index, BES: Binge Eating Scale, AUDIT: Alcohol Use Disorders Identification Test, FTND: Fagerstrom Test for Nicotine Dependence, C-VAT: Clinical Video Game Addiction Test 2.0, CPGI: Canadian Problem Gambling Index.

| Groups | HC (n=178) | BE-only (n=32) | BE + AD (n=30) | F / $\chi^2$ |
| --- | --- | --- | --- | --- |
| Gender (Female) | 112 (62.92%) | 29 (90.63%) | 20 (66.67%) | 9.430** |
| Age (year) | 28.62 (9.78) | 30.03 (8.51) | 28.43 (9.02) | 0.319 |
| BMI (kg/m<sup>2</sup>) | 22.06 (3.29) | 23.70 (3.96) | 22.81 (3.34) | 3.484*<sup>a</sup> |
| BES | 7.35 (4.79) | 24.75 (6.88) | 26.73 (6.37) | 275.200***<sup>ab</sup> |
| AUDIT | 4.90 (3.51) | 4.78 (4.11) | 11.6 (9.39) | 26.950***<sup>bc</sup> |
| FTND | 0.17 (0.74) | 0.19 (0.78) | 1.00 (1.91) | 9.492***<sup>bc</sup> |
| C-VAT | 0.81 (1.35) | 0.69 (1.53) | 5.80 (4.01) | 90.490***<sup>bc</sup> |
| CPGI | 0.22 (0.80) | 0.38 (0.94) | 3.90 (5.18) | 45.220***<sup>bc</sup> |

\*: p<.05, \*\*: p<.01, \*\*\*: p<.001.
a: p < 0.05 for HC vs. BE-only. b: p < 0.05 for HC vs. BE+AD, c: p< 0.05 for BE-only vs. BE+AD

As indicated in Table 1, a significant difference was observed in the gender ratio across the HC, BE-only, and BE+AD group (χ^2^(2)=9.430, p=0.009), while there was no significant difference in age (F(2)=0.319, p=0.727). There was also a statistically significant difference in body mass index (BMI) among the three groups (F(2)=3.484, p=0.032), and the post-hoc analysis revealed that the BMI of the BE-only group was significantly higher than that of the HC group (p=0.031). The effects of gender and BMI were controlled in the ANCOVA for the remaining data analyses.

### 3.2 Binge-eating and Addictions

#### Binge-eating (BES)

The BES scores showed a profound increase in both binge-eating groups compared to the HC group, as shown in Table 1. ANCOVA and post-hoc tests revealed significant differences between the HC and two binge-eating groups (i.e., BE-only and BE+AD) (F=280.02, p < .001). The two binge-eating groups were not significantly different (p = 0.178).

#### Addiction scales

Descriptive statistics and the ANCOVA results regarding addiction scales (alcohol use, nicotine dependence, gaming, gambling) are presented in Table 1. Only the BE+AD groups differed significantly from both the HC and the BE-only groups in all addiction scales. The BE-only group was not significantly different from the HC group in any of the addiction scales (all p > 0.417, post-hoc Tukey’s test).

### 3.3 Cognitive characteristics

Descriptive statistics and the results of ANCOVA regarding cognitive characteristics (behavioral activation/inhibition, impulsivity, self-control, emotion regulation) are presented in Table 2.

**Table 2.** ANCOVA results of psychometric variables. BIS/BAS: Behavioral Inhibition/Activation System Scale. BIS-11: Barratt Impulsiveness Scale-11, BSCS: Brief Self-Control Scale, ERQ: Emotion Regulation Questionnaire, PHQ-9: Patient Health Questionnaire-9, PSS: Perceived Stress Scale, STAI-X: State/Trait Anxiety Inventory-X.

|  | HC | BE-only | BE+AD |  | Post-hoc P-values |  |  |
| --- | --- | --- | --- | --- | --- | --- | --- |
|  | Mean (SD) |  |  | F | HC vs.<br>BE-only | HC vs.<br>BE + AD | BE-only<br>vs. BE +<br>AD |
| BIS/BAS:<br>Inhibition | 19.63 (3.88) | 22.53 (3.08) | 22.30 (3.97) | 12.008*** | **.002** | **.003** | >.99 |
| BIS/BAS:<br>Drive | 11.02 (1.97) | 11.81 (2.58) | 10.57 (2.80) | 2.291 | .135 | .805 | .122 |
| BIS/BAS:<br>Fun | 10.17 (2.35) | 11.06 (2.37) | 11.03 (2.99) | 3.015 | .127 | .142 | >.99 |
| BIS/BAS:<br>Reward | 14.86 (2.49) | 16.06 (2.95) | 15.80 (2.40) | 3.866 | .059 | .215 | .877 |
| BIS-11 | 57.56 (9.78) | 63.16 (10.44) | 74.17 (14.31) | 29.746*** | **.017** | **<.001** | **<.001** |
| BSCS | 37.80 (7.93) | 33.31 (8.29) | 26.30 (7.62) | 25.830*** | **.015** | **<.001** | **.002** |
| ERQ:<br>Reappraisal | 29.06 (6.06) | 27.97 (6.36) | 24.37 (7.24) | 5.624* | .369 | **.002** | .250 |
| ERQ:<br>Suppression | 16.43 (5.03) | 15.44 (5.99) | 17.53 (5.27) | 1.118 | 0.855 | 0.584 | 0.469 |
| PHQ-9 | 4.19 (4.17) | 6.16 (5.20) | 10.73 (6.88) | 25.210*** | 0.102 | **<.001** | **.002** |
| PSS | 16.47 (6.70) | 21.69 (7.08) | 23.97 (6.21) | 21.393*** | **<.001** | **<.001** | 0.438 |
| STAI-X:<br>State | 41.28 (12.04) | 50.31 (13.61) | 55.30 (12.58) | 21.025*** | **<.001** | **<.001** | 0.443 |
| STAI-X:<br>Trait | 42.34 (11.12) | 50.41 (10.23) | 56.50 (10.60) | 25.608*** | **<.001** | **<.001** | 0.155 |

\*p<.05, \*\*p<.01, \*\*\*p<.001 after Bonferroni correction

#### Behavioral Inhibition/Activation Systems (BIS/BAS)

In the BIS/BAS, only the BIS/BAS-Inhibition (behavioral inhibition) showed a significant group difference, while other three subfactors of BAS (drive, fun-seeking, reward responsiveness) did not exhibit significant differences after Bonferroni correction (BIS: F=12.01, p<0.001. Drive: F=2.29, p>0.99. Fun: F=3.02, p=0.611. Reward: F=3.87, p=0.268). Post-hoc tests revealed that behavioral inhibition scores were significantly higher in both the BE-only and the BE+AD groups compared to the HC group (p=0.002 and p=0.003, respectively). Still, there was no significant difference between the two binge-eating groups (p=0.997).

#### Impulsivity (BIS-11)

Average BIS-11 scores showed a stepwise increase across three groups (Mean ± SD: 57.56 ± 9.78, 63.16 ± 10.44, and 74.17 ± 14.31 for the HC, BE-only, and BE+AD groups, respectively). ANCOVA and a post-hoc test revealed a significant difference in BIS-11 among all pairs of the groups (F=29.746, p < 0.001).

#### Self-Control (BSCS)

Average BSCS scores demonstrated a stepwise decrease across three groups (Mean ± SD: 37.80 ± 7.93, 33.31 ± 8.29, and 26.30 ± 7.62 for the HC, BE-only, and BE+AD groups, respectively). ANCOVA and post-hoc tests revealed a significant difference among all pairs of the groups (F=25.830, p<0.001).

#### Emotion regulation (ERQ)

In the ERQ, a significant difference among groups was observed only in the reappraisal factor (F=5.624, p<0.05). Post-hoc analysis revealed that the HC group had significantly higher scores than the BE+AD group (p=0.002).

### 3.4 Mental health characteristics

Descriptive statistics and the results of ANCOVA regarding mental health characteristics (depression, stress, anxiety) are presented in Table 2.

#### Depression (PHQ-9)

The PHQ-9 scores showed a significant group difference in ANCOVA (F=25.210, p<0.001). Post-hoc tests revealed significant differences between the BE+AD groups vs. the other two groups: p < 0.001 for BE+AD > HC, p=0.002 for BE+AD > BE-only. The difference between the HC and the BE-only was insignificant (p = 0.102).

#### Stress (PSS)

The PSS scores showed a profound increase in both binge-eating groups when compared to the HC group (F = 21.393, p < 0.001). Post-hoc tests revealed a significant difference between the HC group and both binge-eating groups (all p<0.001), but no significant difference was observed between the BE-only and BE+AD groups (p=0.438).

#### State and Trait Anxiety (STAI-X)

Both state and trait anxiety scores showed a significant group difference in ANCOVA. In post-hoc tests, both binge-eating groups had significantly increased state and trait anxiety than the HC group (all p < 0.001), while exhibited no significant difference between the BE-only and BE+AD groups (p=0.443 and p=0.155 for state and trait anxiety, respectively).

## 4 Discussion

In ANCOVA controlled for the effects of gender and BMI, we observed significant group differences across the HC, BE-only, and BE+AD groups in the following psychometric scales: BIS/BAS-Inhibition, BIS-11, BSCS, ERQ-Reappraisal, PHQ, PSS, and STAI-X (both state and trait anxiety).

First, there were significant effects of binge-eating in BIS/BAS-Inhibition, PSS, and STAI-X (state and trait anxiety). In these scales, there were no group differences related to the comorbid addiction status. Thus, both binge-eating groups showed similar increases in behavioral inhibition (in the motivation system), perceived stress, and anxiety when compared to the HC groups. Second, there was a significant increase in BIS-11 (impulsivity) and a decrease in BSCS (self-control) stepwise across the HC vs. BE-only vs. BE+AD groups. These factors might be associated with more severe psychopathological alterations in BED with comorbid addiction. Third, we observed unique alterations in the BE+AD groups for PHQ (depression) and ERQ-Reappraisal. Only the BE+AD groups showed significantly stronger depression symptoms and impaired cognitive reappraisal in emotion regulation when compared to the HC group. The BE-only group did not significantly differ from the HC group in these mood-related symptoms. It may result from the interplay of binge eating and addiction symptoms or simply the consequence of the addictive disorders. For example, co-occurrence of mood disorders can be more frequent in BED patients with substance use disorders (Becker and Grilo, 2015).

The prevalence of binge eating (or BED) was much higher in the present study (total of 62 out of 312 subjects, resulting in 19.9%) than the previously known prevalence rate of BED in the general population. In the present study, most participants were young females (as described in the Methods and in Table 1) who had an elevated risk of BED. As we recruited voluntary participants via online advertisement, it might draw much more attention among the participants who are concerned with eating problems or addictive behaviors. It might also be associated with the potential bias in gender and age in the samples. Finally, the present study screened individuals with binge-eating problems using the self-reported screening tool, i.e., BES, which might be less accurate than the structured clinical interview. A study validating BES for diagnosing BED in the female population in Portugal demonstrated remarkable sensitivity and specificity (81.8% and 97.8%, respectively), but such validation was not rigorously conducted with Korean populations. Therefore, the binge-eating groups in this study might include individuals with the sub-clinical level of binge-eating behavior. Still, it will be meaningful to assess the effect of comorbid addictions in the binge-eating population in the present study, as our binge-eating subgroups (BE+AD vs. BE-only) were based on the same screening criteria and the effects of other demographic variables (age and BMI) were controlled with ANCOVA.

In contrast to previous studies, the present study assessed frequent substance addictions (alcohol and nicotine) and behavioral addictions (gambling and video games) together to identify the comorbid addiction group. Among 30 BE+AD subjects, 17 had substance addiction (16 for alcohol, one for nicotine), but the remaining 13 had only behavioral addictions. These binge-eating individuals with co-occurring behavioral addictions might have been ignored so far in previous studies of comorbidity of addiction in the binge-eating population. It might be interesting to contrast the effects of substance addiction vs. behavioral addiction in the binge-eating population, but we abandoned it due to the limited sample size and the heterogeneity of the BE+AD group in the present study (some participants also exhibited both types of addiction). However, the present study design is highly scalable to a very large population when combining with online survey tools. A further study with a larger sample size can provide more concrete evidence and more detailed sub-group comparisons (e.g., the effect for the types and the numbers of comorbid addictions).

There are a few other limitations in the present study. First, screening of the addictive disorders was also based on self-reported clinical questionnaires. Thus, it might be less accurate and inflated the prevalence of each addiction. However, the self-report-based study has its advantage in the scalability as noted above. Alternatively, the following studies utilizing both clinical interviews and the self-reported clinical scales can prove the reliability of the present findings. Second, the present study cannot directly suggest the causal relationship between the study variables. There can also be other mediating factors between the observed relationship between the psychometric properties and the group status (binge eating and addiction) in this study. Structured models on the theoretical basis can be constructed and validated with additional data in the future.

Our study is a systemic evaluation using a set of relevant psychometric scales encompassing a range of cognitive functions and mental health characteristics that are commonly implicated in binge eating and addictive disorders. Increased impulsivity and decreased self-control can be major predisposing factors for binge-eating and addictive behaviors, which might result in impaired emotion regulation and severe depression symptoms in the comorbid condition. We found robust group differences after a stringent Bonferroni correction for multiple comparisons, so the findings will likely be replicated in future studies with a larger sample. Our findings might be helpful to construct a more extensive theoretical framework for understanding the nature of binge-eating and comorbid addictive disorders, particularly by demonstrating relevant cognitive constructs for further investigation of the shared neural mechanisms of these psychiatric conditions and the potential treatment targets in cognitive-behavioral therapy.

## Data Availability

All data produced in the present study are available upon reasonable request to the authors.

## 5 Conflict of Interest

We have no conflicts of interest to disclose.

## 6 Author Contributions

“J.Jeong: Writing – original draft, Conceptualization, Data curation, Formal analysis, Investigation, Methodology; J.Jang: Writing – review & editing, Validation; G.Jeon: Writing – review & editing, Investigation; K.Baek: Writing – original draft, review& editing, Conceptualization, Methodology, Supervision, Validation.”

## 7 Funding

This study was partly supported by the National Research Foundation of Korea (NRF-2021R1F1A1063968), Institute of Information & communications Technology Planning & Evaluation (IITP) under the Artificial Intelligence Convergence Innovation Human Resources Development (IITP-2023-RS-2023-00254177) grant funded by the Korea government (MSIT), and Ministry of Education of South Korea (the BK21 Four program, Korean Southeast Center for the 4th Industrial Revolution Leader Education, Pusan National University).

## 8 Acknowledgments

## Notes

### Competing Interest Statement

The authors have declared no competing interest.

### Author Declarations

The Institutional Review Board (IRB) of Pusan National University gave ethical approval for this work.

